# MedEvalarena: A Self-Generated, Peer-Judged Benchmark for Medical Reasoning

**DOI:** 10.64898/2026.01.27.26344905

**Authors:** Preethi Prem, Kie Shidara, Vikasini Kuppa, Esmé Wheeler, Feng Liu, Ahmed Alaa, Danilo Bernardo

## Abstract

Large Language Models (LLMs) demonstrate strong performance at medical specialty board multiple-choice question (MCQ) answering, however, underperform in more complex medical reasoning scenarios. This gap indicates a need for improving both LLM medical reasoning and evaluation paradigms. We introduce MedEvalArena, a framework in which LLMs engage in a symmetric round-robin format. Each model generates challenging board-style medical MCQs, then serves in an ensemble LLM-as-judge bench to adjudicate validity of generated questions, and finally completes the validated exam as an examinee. We compared performance of leading LLMs across the OpenAI, Grok, Gemini, Claude, Kimi, and DeepSeek families on both question generation validity and exam-taking performance. Across frontier models, we observe no statistically significant differences in exam-taking performance, suggesting convergence in current medical reasoning ability across frontier LLMs for question-answering. We observed higher question validity rates in questions generated by OpenAI, Gemini, and Claude frontier models compared to Kimi, Grok, and DeepSeek models. When jointly considering accuracy and inference cost, multiple frontier models lie on the Pareto frontier with no single model dominating across both dimensions. MedEvalArena provides a dynamic and scalable framework for benchmarking LLM medical reasoning.

## 1 Introduction

The performance of Large Language Models (LLMs) in medical reasoning tasks has quickly progressed ^1^, now matching or exceeding human physician performance on standardized medical exams such as the United States Medical Licensing Examination (USMLE) Step exams ^2–4^. However, as LLMs continue to improve, their performance remains limited in medical benchmarks requiring open-ended reasoning and future planning ^5–9^, demonstrating the need for more robust LLM medical reasoning and evaluation frameworks.

Conventional medical reasoning evaluation relies on benchmarks which requires academic peer review and validation ^10^. However, LLM reasoning capabilities are improving rapidly, driving a continuous need for novel and more challenging benchmarks of reasoning capabilities ^11^. This introduces significant lag between the abilities of frontier LLMs and our understanding of their medical reasoning competence. Addressing these limitations motivates the need for dynamism in evaluation frameworks that are able to adapt alongside LLMs, specifically evaluation pipelines that enable rapid benchmark re-instantiation to facilitate a tighter feedback-loops for LLM improvement.

To mitigate benchmark saturation, dataset contamination, and to keep pace with rapid model development, recent work has introduced LLM benchmarks that can be updated with human guidance. Approaches such as DynaBench ^12^ and LiveBench ^13^ are capable of intermittent updates of assessments, with human oversight of task curation. BenchBuilder operationalizes updating at scale via automatic generation of a sizeable, diverse bench of tasks at low cost ^14^. More recently, Autobench incorporates the LLM-as-judge paradigm into dynamic benchmark generation, utilizing a round-robin framework where models generate questions, complete questions, and evaluate outputs through peer-assessment ^15^.

Motivated by the potential of these approaches to assess and advance LLM performance in medical reasoning, we present MedEvalArena, a dynamic framework for evaluating the medical reasoning capabilities of LLMs through peer assessment. In this approach, LLMs participate in a symmetric round-robin design in which each model generates challenging medical questions designed to assess other models’ reasoning capabilities. We utilize an LLM-as-judge ensemble paradigm to ensure validity of LLM-generated questions across two dimensions, logical coherence and medical accuracy ^16;17^. All models are then evaluated on the complete question set, enabling scalable benchmarking of LLM medical reasoning capabilities.

## 2 Methods

We construct our MedEvalArena in three stages (Figure 1): (i) question generation by LLMs acting as question generators, (ii) LLMs serving on judge ensemble that assess item validity, and (iii) a round–robin evaluation in which multiple LLMs take all questions as examinees.

**Figure 1.**
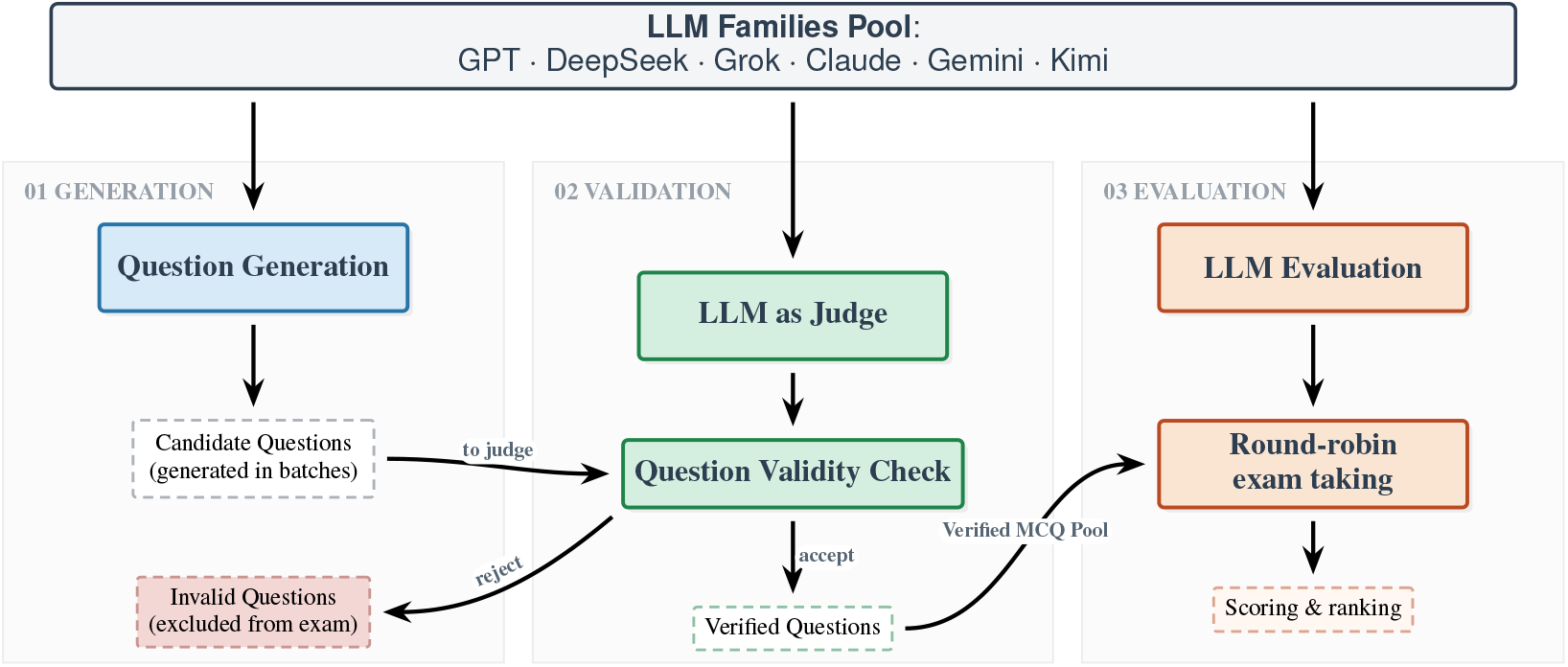
Round-robin LLM evaluation scheme. Each of six LLM families first generates questions while the LLM-as-judge ensemble (generation and validation phases), consisting of the same six judges adjudicates the validity of the generated questions. The resulting set of valid questions is then used as a shared benchmark: every model answers every question, and aggregate performance across the entire exam is used to compare models.

### 2.1 Stages One and Two: Round-robin Question Generation and Peer Judging

In the first stage, each generator model *g* generates board-level medical multiple–choice questions. For each *g*, we specified a total of 50 questions with topic distribution determined by specialty representation of the US physician workforce as published by the Association of American Medical Colleges ^18^. This provides a population-based representation of generalist core questions from internal medicine and family medicine and specialty board questions.

Subspecialty names were harmonized to match the American Board of Medical Specialties (ABMS) specialty names. The union of subspecialty topics within internal medicine and family medicine were obtained from American Board of Family Medicine and American Board of Internal Medicine Board topic distribution websites ^19;20^. The topic distribution is provided in Supplementary Table 1. The generator is instructed, via a structured system prompt, to produce extremely difficult medical board–style questions with exactly one best answer and an accompanying explanation for that answer. Each LLM model is queried once per question with this specification, using a low sampling temperature (*T* = 0) in order to favor reproducibility.

To filter out clinically inaccurate or logically incoherent items, we employ an ensemble LLM-as-judge phase that reviews each question–answer pair. The judge ensemble receives, for every question, the question stem, the full list of answer options, the designated correct answer, and the explanation produced by the generator, together with a detailed rubric describing the desired evaluation criteria. For each question, each judge in the judge ensemble is prompted to return a structured assessment in which it provides (i) a free–text analytic rationale, (ii) an integer medical accuracy score on a five–point ordinal scale, (iii) a binary indicator of logical validity specifying whether the item is well–posed and has a defensible unique answer, and (iv) a categorical label that summarizes the primary failure mode when logical validity does not hold, specifically one of the following: contradiction, miskeyed answer, multiple defensible answers, or underspecification. Medical accuracy score in rated on the following scale: 1 = Dangerous/False: clear hallucinations or advice that plausibly causes harm; strongly contradicts standard care. 2 = Inaccurate: major medical errors or clearly outdated guidance that would mislead a learner. 3 = Minor issues: mostly correct but missing important details, uses nonstandard terminology, or is clinically unrealistic in a non-critical way. 4 = Accurate: consistent with current standard care/guidelines; clinically sensible. 5 = Gold standard: fully accurate, precise, realistic, and board-relevant.

For questions to be included for the subsequent primary outcome evaluation phase, the question is kept only if a strict majority of judges rate it logically valid and the strict majority of judges assign a medical-accuracy score of *τ* ≥ 4. Question generators were prompted to generate valid questions until the specified AAMC/ABMS distribution of 50 questions per exam was met.

### 2.2 Stage Three: Round–Robin Exam Evaluation

We evaluate each model *a* on all questions produced by each generator model *g*. For each question, the taker is given the stem and five answer options and instructed to output exactly one option label as its final answer. All taker queries use *T* = 0 for reproducibility. For each (*g, a*) pair, we compute accuracy as the proportion of questions answered correctly. We report mean accuracy for each generator–taker pair.

### 2.3 Statistical Analysis

We evaluated for differences in LLM question generation validity rates with pairwise two-sample proportion tests. We performed leave-one-out cross-validation (LOOC) agreement of individual judges across all questions to assess individual judge agreement against the ensemble. Each judge is held out in turn and compared against the consensus formed by the remaining five judges (N=5). To assess logical validity, agreement is measured as accuracy with the ensemble’s binary pass/fail decision. For medical accuracy score, agreement is measured as exact match with the ensemble’s median score. 95% confidence intervals were generated via boostrap permutation with 5000 bootstraps. We tested for differences in item-level question accuracy across LLMs. Using R `lme4`, we fit a binomial-logit GLMM with LLM identity as a fixed effect and random intercepts for question-within-exam ^21;22^. We tested the LLM effect via a likelihood-ratio comparison to a null model ^23^. All statistical comparisons were adjusted for multiple comparisons with Benjamini–Hochberg procedure.

### 2.4 Code and Dataset Availability

The MedEvalArena dataset and analysis code are available at https://github.com/bernardolab/medEvalArena.

## 3 Results

We included LLMs for exam generation and LLM-as-judge that were the leading models from the top 6 frontier labs from artificialanalysis.ai rankings with model cutoff date of Nov 20, 2025^24^. This resulted in six exam-generators including: OpenAI gpt-5.1-2025-11-13^25^, Anthropic claude-opus-4.1^26^, Google gemini-3-pro-preview ^27^, deepseek-v3.2^28^, XAI grok-4-0709^29^, and Kimi kimi-k2-thinking ^30^. These six models subsequently also served in the LLM-as-judge ensemble.

Each exam-generator was used to create a single 50 question exam per generator, following topic distribution weighted by the U.S. physician specialty distribution reported by the AAMC ^18^, yielding a total of 300 questions containing both generalist and specialty board questions. Topic distribution is shown in Supplemental Information 1. To assess question validity we evaluated each generated MCQ under an LLM-as-judge paradigm using strict rating criteria for medical accuracy and logically coherence. Questions were deemed valid if and only if the strict majority of judges rated the question as logically coherent and if the majority of LLM judges rated each question as medical accurate with score greater than or equal to 4. Across all models, gpt-5.1 achieved the highest proportion of generated questions passing as valid with 94.8% and deepseek-v3.2 had the lowest proportion passing as valid with 46.0%. Pairwise proportion tests indicate no detectable difference between GPT-5.1, Gemini, and Claude, while these models outperformed DeepSeek, Grok, and Kimi models (Figure 2b).

**Figure 2.**
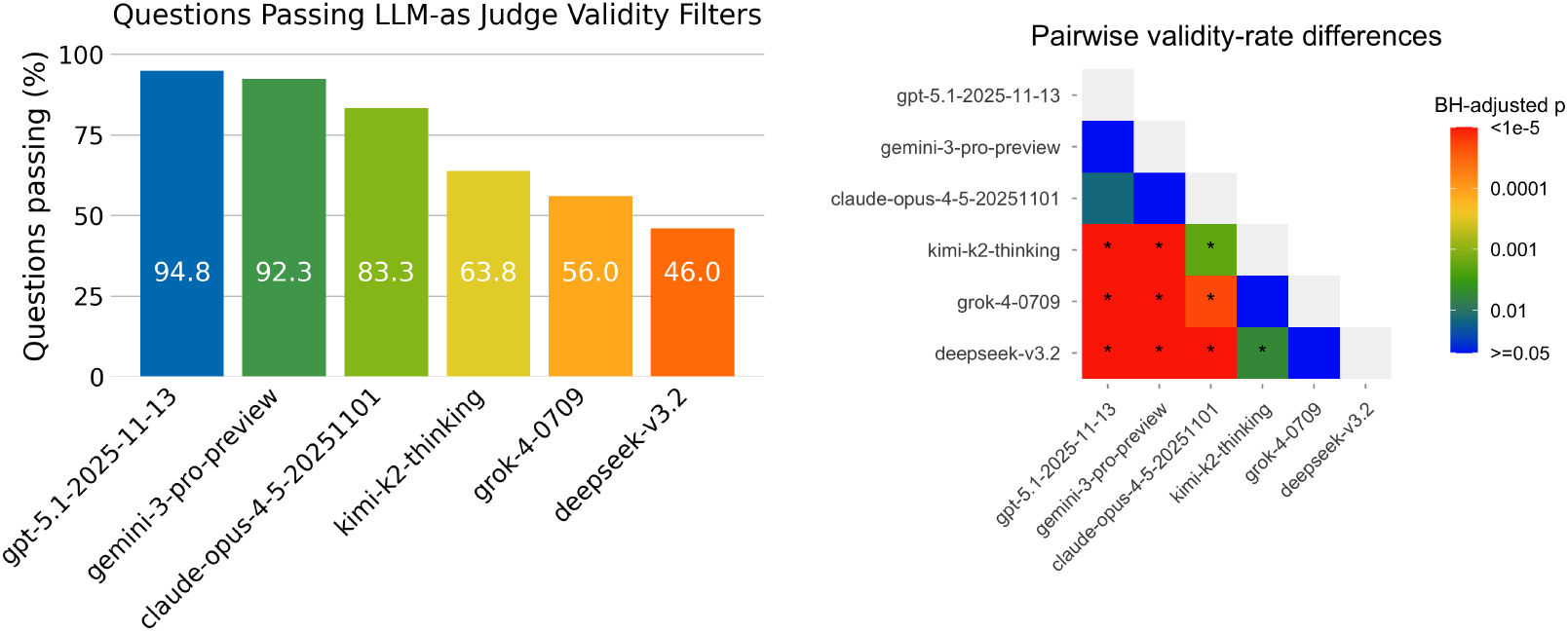
(a) Bar plot demonstrating question validity rates of each question generator. Across generators, gpt-5.1-2025-11-13 produces questions with the highest percentage of validity (94.8.%). (b) Heatmap of Benjamini–Hochberg (BH)–adjusted *p*-values from pairwise two-sample proportion tests comparing validity rates from (a) across pairs of generators. Color encodes − log_10_(BH-adjusted *p*), with red indicating stronger evidence for a difference between the generator pairs. Asterisks mark comparisons with BH-adjusted *p <* 0.01.

Next, we use leave-one-out cross-validation (LOOC) to evaluate each judge by holding it out of the judge ensemble, forming a 5-judge consensus (N=5) from the remaining models, and measuring agreement with that consensus over all questions. For logical validity (Figure 3a), agreement is reported as accuracy, defined as the fraction of items on which the held-out judge matches the ensemble’s logical validity binary pass or fail decision. Accuracies range from approximately 0.72 to 0.87, indicating moderate consistency with the ensemble. For medical accuracy score (Figure 3b), agreement is the fraction of items for which the held-out judge exactly matches the consensus median score. Agreement rates ranged between 0.64 to 0.79 reflecting broadly consistent scoring across judges with mild disagreement around the ensemble median. Overall, we did not observe any outlier judges for both logical validity and medical accuracy evaluation dimensions.

**Figure 3.**
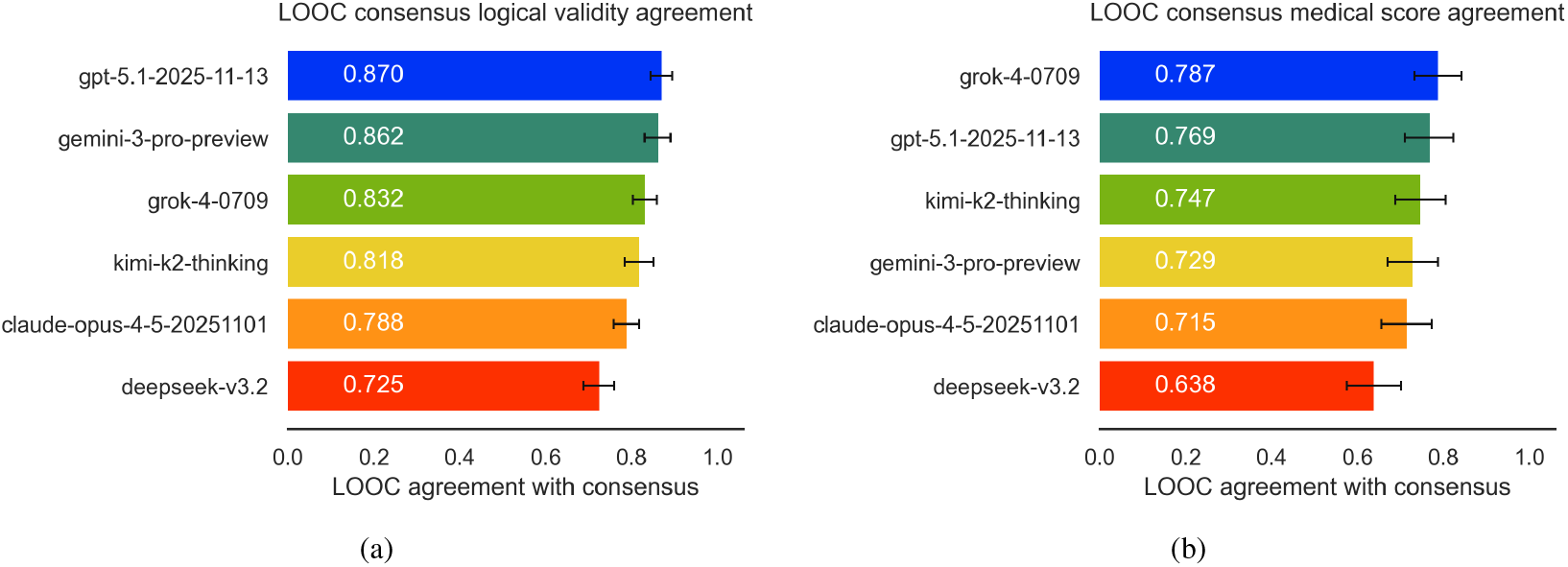
Leave-one-out cross-validation (LOOC) agreement of individual judges across all questions. (a) Logical validity: agreement is measured as accuracy with the ensemble’s binary pass/fail decision. (b) Medical accuracy score: agreement is measured as exact match with the ensemble’s median score. 95% confidence intervals are shown.

In the evaluation phase, the accuracy of each exam-taker was assessed on the full set of validated questions (N=300). Visualization of generator–by–taker matrix (Figures 4 and 5) reveals how well models perform across generated exams. While the generator–by–taker matrix shows modest variation across model pairs and across exam sources for frontier LLMs, none of these differences were statistically significant. In the question generation phase, claude-opus-4.5 generated the most challenging exams (average accuracy of 84.3%) whereas gemini-3-pro-preview generated the least challenging exams (average accuracy of 96.3%). In the exam-taking stage, claude-opus-4.5 performed best with the highest answer accuracy (91.7%), followed by gpt-5.1-2025-11-13 (88.7%) and deepseek-v3.2 (88.3%). Model performance did not differ significantly, as assessed by a binomial-logit generalized linear mixed model (GLMM).

**Figure 4.**
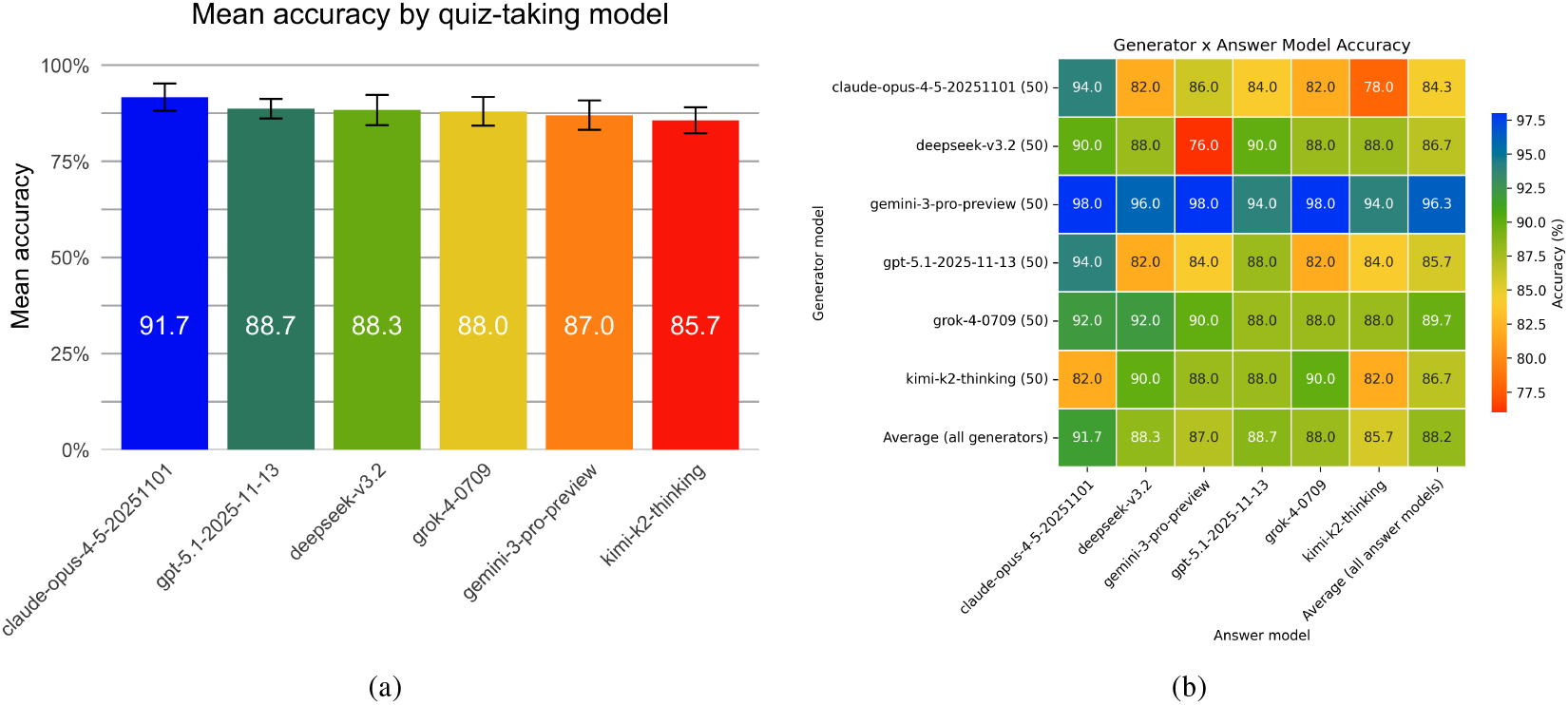
(a) Exam model accuracy (b) Generator × answer model accuracy heatmap. Each row corresponds to the model used to generate questions (generator), and each column to the model used to answer those questions (answer model). Cells show the mean accuracy for each generator–answer pair, expressed as the percentage of questions answered correctly and averaged across all exams (one 50 question exam per generator). The rightmost column reports, for each generator, the average accuracy across all answer models. The bottom row reports, for each answer model, the average accuracy across all generators. The bottom-right cell gives the overall mean accuracy across all model pairs (88.2%).

**Figure 5.**
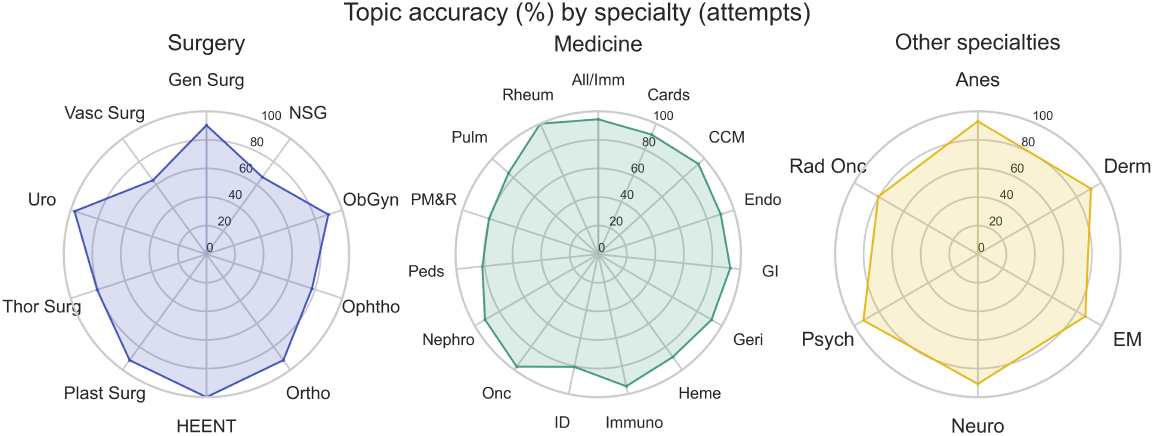
Radar plots depicting percentage accuracy of all six exam-takers across 31 topics, grouped by specialty into surgical, medical, and other specialties. Accuracy was calculated from 300 validated questions, with mean correctness shown per topic, where greater radial extent indicates higher accuracy.

Additionally, we compared exam-taker accuracy among medical specialties and found that the six LLMs as a whole demonstrated the lowest performance in surgical topics, particularly vascular surgery (63.9%) and neurosurgery (66.7%), while exceeding 80% accuracy in all non-surgical specialties and reaching 100% accuracy in rheumatology and musculoskeletal topics (Figure 5). These findings were unchanged when accounting for unequal numbers of questions per topic. To determine whether question difficulty differed by specialty, we conducted a global permutation ANOVA on mean question correctness and found no significant differences across specialties.

Next, we evaluated inference cost versus accuracy trade-offs, including weaker reasoning models from Grok, Claude, Gemini, and GPT families. While accuracy differed modestly across models (76.0–91.7%), per-exam cost varied by nearly two orders of magnitude (0.186–9.27; Fig. 6). The resulting Pareto frontier consisted of `grok-4-1-fast-reasoning` (86.3%, 0.1855), `deepseek-v3.2` (88.3%, 0.42), `gpt-5.1-2025-11-13` (88.7%, 1.86), and `claude-opus-4-5-20251101` (91.7%, 2.15), with all other models dominated with higher cost at equal or lower accuracy.

**Figure 6.**
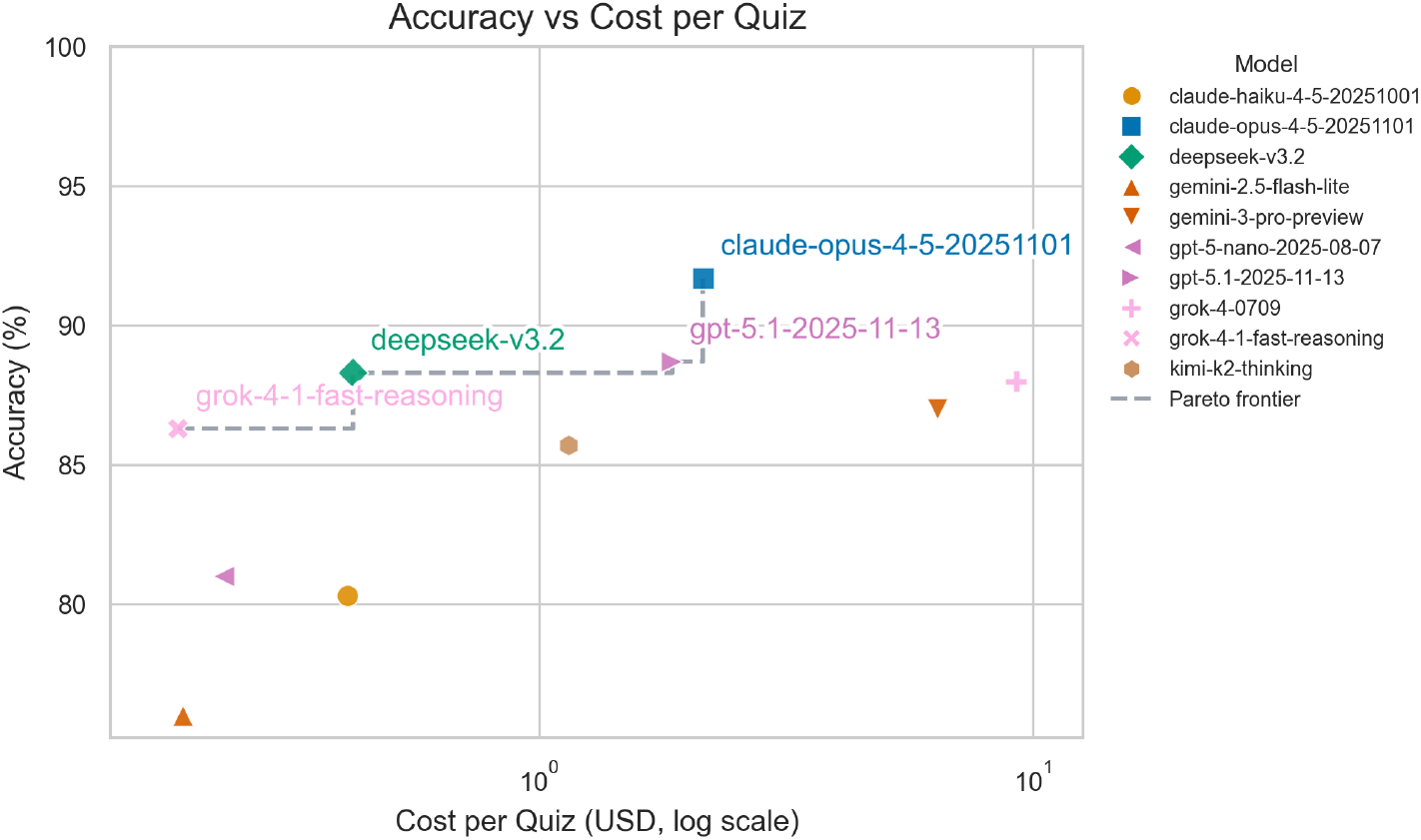
Accuracy versus cost per exam scatter plot. Cost-per-exam is the cost for an LLM to complete the entire 300 question exam. Accuracy differed only modestly across models, however, per-exam cost varied by two orders of magnitude. The Pareto frontier (gray dashed lined) consisted of `grok-4-1-fast-reasoning` (accuracy 86.3%, cost per exam $0.19), `deepseek-v3.2` (88.3%, 0.42), `gpt-5.1-2025-11-13` (88.7%, 1.86), and `claude-opus-4-5-20251101` (91.7%, 2.15).

In a secondary sensitivity analysis, we evaluated the effect of varying minimum medical accuracy score for exam inclusion (Fig 7). We found that only 4 questions (1.3%) had medical accuracy score of 4, and the remainder (98.7%) had medical accuracy of 5. Given the small number of questions with medical accuracy score of 4, we did not assess for effect of varying question medical accuracy score on model accuracy.

**Figure 7.**
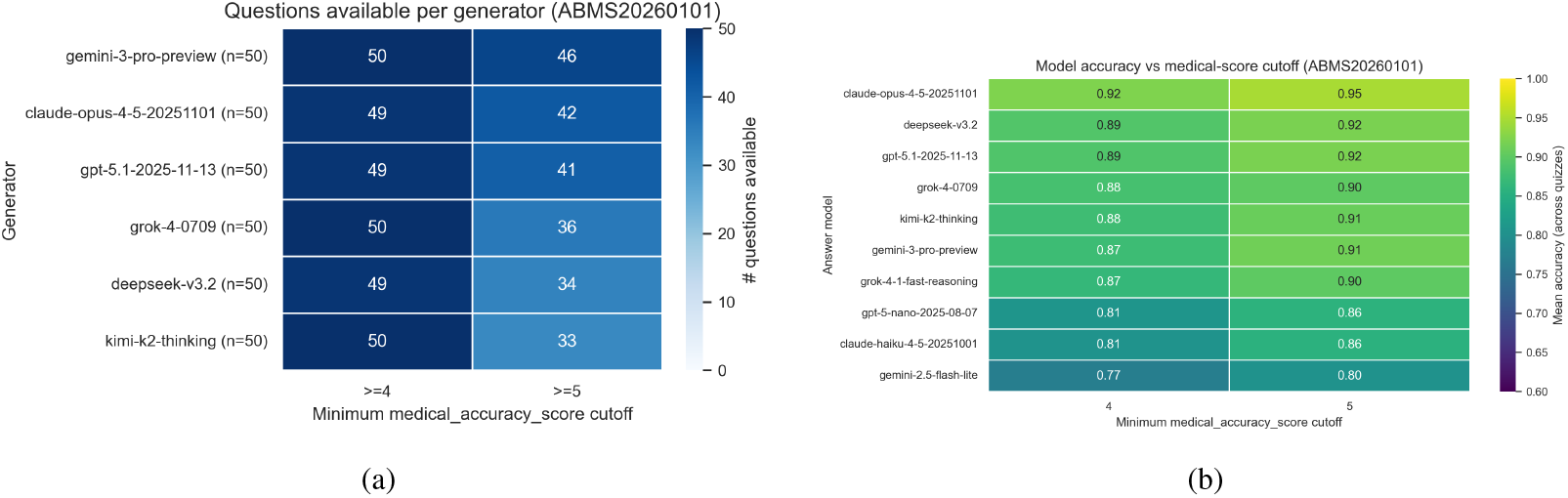
Sensitivity analysis. (a) Demonstrates how many questions per generator achieved different medical accuracy cut-offs (4 or 5). (b) Demonstrates accuracy per generator when utilizing only questions that achieved minimum medical score accuracies of at least 4 or 5.

## 4 Discussion

We utilized symmetric round robin peer-assessment to assess medical reasoning of LLMs and found no significant difference in the MedEvalArena exam-taking accuracy of frontier models. This convergence suggests that frontier models have reached parity on competency on medical-board style multiple choice reasoning tasks. Notably, we found differences in generator validity rates, suggesting that reasoning processes supporting question generation capability may be a distinguishing factor in medical reasoning performance. Also, we found that while the spread in model accuracy was modest, per-exam inference cost differed by nearly two orders of magnitude, producing a Pareto frontier amongst frontier models.

The finding that question generation quality separates models more than exam-taking accuracy supports the view that question-answering and question-generation involve distinct cognitive and reasoning capabilities ^31^. Generation requires not only medical knowledge, but also the ability to pose well-specified clinical vignettes with logical coherence ^32^, necessitating a uniquely defensible single best answer, and avoiding underspecification ^33^. The reasoning processes supporting these facets of question generation are relatively more recursive and nonlinear than question answering ^34^, and may be more difficult for LLMs due to their relative deficiencies in planning ^35^. Question-generating capability reflects underlying medical reasoning and specification ability ^36^ and thus is relevant to clinical reliability, as underspecification or internal inconsistency may lead to ambiguous or misleading medical guidance. Nonetheless, we acknowledge that exam-taking performance and question-generation ability are only proxy measures of real-world clinical reasoning.

Our approach extends prior dynamic benchmarking efforts ^12;13^. In this respect, MedEvalArena is similar to the reciprocal, round-robin evaluation approach in Autobench ^15^, while being focused on medical reasoning. Our also approach differs from Autobench in that due to the multiple choice nature of MedEvalArena, we utilized LLM-as-judge phase immediately after question-generation to validate question coherence rather than grade responses.

We acknowledge several limitations of this work. First, statistical power to detect differences in LLM exam-taking performance may be limited by the limited number of questions generated. Nonetheless, the absolute differences separating frontier model accuracies were modest. Second, LLM as judge paradigm may introduce potential bias such as shared training artifacts between LLM families. The potential for biases motivates development of stronger adjudication protocols which may include larger judge ensembles. Human rating of question validity may be helpful, however, due to the scalable and potentially rapidly evolving nature of our approach, human oversight may be partial or limited. In addition, additional analyses including calibration and explanation faithfulness are warranted. Lastly, although MedEvalArena is designed to support dynamic regeneration of updated evaluations, this work evaluates a single snapshot generated with a fixed model roster. The full pipeline is provided to facilitate future MedEvalArena instantiations.

## Data Availability

All data produced are available online at https://github.com/bernardolab/MedEvalArena

## 4.1 Supplementary Tables

**Supplementary Table 1:** AAMC/ABMS Population Based Question Distribution.

| Specialty | Number |
| --- | --- |
| Pediatrics | 5 |
| Emergency Medicine | 3 |
| Obstetrics and Gynecology | 3 |
| Anesthesiology | 2 |
| Psychiatry | 3 |
| General Surgery | 2 |
| Ophthalmology | 1 |
| Orthopedic Surgery | 1 |
| Critical Care Medicine | 1 |
| Neurology | 3 |
| Dermatology | 1 |
| Urology | 1 |
| Otolaryngology | 1 |
| Physical Medicine and Rehabilitation | 1 |
| Plastic Surgery | 1 |
| Neurological Surgery | 1 |
| Radiation Oncology | 1 |
| Thoracic Surgery | 1 |
| Vascular Surgery | 1 |
| Cardiovascular | 2 |
| Endocrine | 2 |
| Gastrointestinal | 2 |
| Pulmonary | 2 |
| Nephrology | 1 |
| Hematology | 1 |
| Immunology | 1 |
| Rheumatology/Musculoskeletal | 1 |
| Infectious Disease | 2 |
| Medical Oncology | 1 |
| Allergy/Immunology | 1 |
| Geriatrics | 1 |

